# A Systematic Review of the Association between the Age of Onset of Spinal-Bulbar Muscular Atrophy (Kennedy’s Disease) and the Length of CAG Repeats in the Androgen Receptor Gene

**DOI:** 10.1101/2023.02.08.23285647

**Authors:** Dante J. Bellai, Mark G. Rae

## Abstract

**Background:** Spinal bulbar muscular atrophy (SBMA) is an X-linked recessive motor neuron disorder which is caused by the presence of ≥ 38 CAG repeats in the androgen receptor gene. Relatively little is known about SBMA, but existing literature indicates a relationship between CAG repeat number and the onset age of some motor symptoms of SBMA. This literature review explored the effect of larger CAG repeats on the age of weakness onset compared to shorter length CAG repeats in male SBMA patients.

**Methods:** Three databases were searched (MEDLINE, SCOPUS, and Web of Science; Oct 2021) along with targeted searches in Cambridge University Press and Annals of Neurology. 514 articles were initially identified, of which 13 were included for qualitative synthesis.

**Results:** Eleven of the thirteen articles identified a statistically significant inverse correlation between CAG repeat length and age of weakness onset in SBMA. Five studies indicated that SBMA patients with fewer CAG repeats (*e.g*. 35-37) had an older age of weakness onset than patients with a greater number (*e.g*. >40) of CAG repeats. The minimum number of CAG repeats associated with weakness was numbered in the mid-to-late thirties.

**Conclusion:** Identification of a relationship between CAG repeat number and weakness may enable earlier detection and intervention for SBMA.

Limitations of this review include the restriction to English-only studies and differences in statistical methodology used in each study. We recommend that future studies use interviews, chart reviews, and standardized scoring methods to reduce effects of retrospective bias on reporting SBMA signs and symptoms.

## Background

Spinal-bulbar muscular atrophy (SBMA), or Kennedy’s disease, is an X-linked recessive motor neuron disorder associated with greater than 38 CAG repeats in the androgen receptor gene (1), that affects approximately 1/40,000 men (although prevalence is greater in certain locations, such as the Vaasa region of Finland) (2). Increased CAG expansion induces toxic polyQ-AR fragments, resulting in the disruption of transcription, mitochondrial function, protein cycling, cell signaling pathways and autophagy (1,3). The average age of onset of SBMA is in the third or fourth decade and, although female carriers display some symptoms of the condition such as distal motor deficits, cramping and/or fasciculations (1), men are much more severely impacted (4) with symptoms that include dysarthria, dysphagia, gynecomastia, cramping, fasciculations and tremor (1).

Generally, SBMA follows a gradual progression with postural tremors of the upper limbs starting in the early thirties (5). In addition to the onset of dysarthria and dysphagia in the forties, lower limb motor deficits also develop, eventually requiring the use of walking aids in the fifth decade (5). Finally, patients become wheelchair-bound by, or during, their sixth decade (5). Patients with bulbar musculature implicated early-stage SBMA often die from recurrent aspiration pneumonia in their fifties (6).

Although it is known that the age of onset of non-motor manifestations of SBMA is independent of CAG repeat length, primary literature does indicate that there is also a clear negative correlation between the age of onset of motor manifestations and CAG repeat number (for review see (4)). Although several studies (5–17) have pondered the relationship between CAG repeat number and the age of the most frequent symptom of SBMA, weakness (18), to date, no systematic review has specifically focused on the possible connection between these two factors. As such, herein we seek to address this deficit by exploring the effect of a greater number of CAG repeats on the age of weakness onset in comparison to a lower number of CAG repeats in male SBMA patients (between the ages of 11 and 83) who did not suffer from other major chronic illnesses.

Given that very little is known about the aetiology of SBMA, probably due to its relatively low prevalence in the population, and that current treatments for SBMA only facilitate symptom management, men with the condition should be identified as soon as possible in order for treatment to begin promptly (4). As such, any addition to the SBMA-associated literature which increases understanding of the disease must be welcomed.

## Objectives

Primary literature was reviewed in order to:

1. Explore whether or not the association between CAG repeat number and age of onset of symptoms of SBMA was, or was not, significant.
2. Determine what minimum and maximum number of CAG repeats was associated with weakness onset in SBMA.

## Materials and Methods

### Study eligibility

Inclusion criteria (see Table 1) were carefully prepared such that they enabled identification of as many articles as possible that addressed the relationship between CAG repeat number and the specific age of onset of weakness in SBMA patients. Studies that were selected must have included at least some male SBMA patients, with female only studies being excluded. Only studies that examined SBMA patients (with or without female carriers) that had no other major health conditions such as a concomitant diagnosis of schizophrenia, were included. Studies that involved patients with health conditions that commonly co-occur with SBMA such as type II diabetes were included. In addition, the selected age profile of participants had to range from 11 to 83, which was the most common age range of study participants that was encountered in our literature search.

**Table 1.** Inclusion and exclusion criteria.

|  | Inclusion | Exclusion |
| --- | --- | --- |
| Intervention | Studies that specifically compared CAG repeat expansion with the onset of muscle weakness in SBMA patients | Studies that addressed activities of daily living but did not specifically compare the onset of weakness with CAG repeat expansion |
| Population | Male SBMA patients only, or mixed male SBMA patients and female carrier patient populations aged 11-83 with SBMA | Studies that were specific to female carriers |
| Participants | Male SBMA patients (with or without female carriers) that had no other major health conditions or that had health conditions that commonly co-occur with SBMA such as Type II Diabetes | SBMA patients (with or without female carriers) that had other major health problems such as Schizophrenia |
| Outcomes | Studies that quantified the number of CAG repeats and compared it with the age at which weakness of any kind (upper limb, lower limb *etc.*) was first reported | Studies that identified the relationship between CAG repeats and age at onset as being defined as the age at which any symptoms appear (not specifically weakness) |
| Date | Given that it is a rare disease, any study may be included no matter how old | N/A |
| Language | Studies written in English | Studies that were not written in English |
| Types of publication | Any kind of primary literature including: RCTs, longitudinal, case-control, mixed methods studies, observational quantitative, *etc.* | Review articles / commentaries / editorial articles |
| Availability | Both freely available full texts and those that had to be purchased | Articles that were not available online |
| Peer reviewed | Studies published in a peer-reviewed journal | Studies not published in a peer-reviewed journal |

Any form of peer-reviewed, primary research study (*e.g*. case control study, longitudinal study, randomized controlled trial, observational quantitative, mixed methods, *etc*.), as long as it was written in English and not conducted using animals, was eligible for inclusion. However, reviews, commentaries and/or editorials were excluded. Both freely available texts as well as those that could only be purchased (7) were included as long as they could be accessed online. Given the rare nature of the disease and the relative dearth of available literature pertaining to it, we did not apply a date range to our searches and as such studies were accepted even if they were several decades old.

### Study Identification

Searches were conducted between September and October 2021, by an independent reviewer, and after searching ‘Spinal Bulbar Muscular Atrophy’ and ‘CAG repeat’ on EBSCO, the most commonly cited database was PubMed (*a.k.a*. MEDLINE). PubMed was searched using the following combined terms that were designed to address three concepts: “spinal bulbar muscular atrophy”, “age of onset of muscle weakness”, and “CAG repeat”, which yielded 116 results. Additional searches were made of the SCOPUS and Web of Science databases, utilizing variations of the same three concepts, which generated 390, and three results, respectively. Individual searches were also conducted in two journals: firstly, Cambridge University Press where the abbreviated term “SBMA” was used (yielding two results), and the Annals of Neurology: Wiley Online Library where the search terms for a specific author of interest (“Manabu Doyu”) were used, in addition to the disease of interest (“spinal bulbar muscular atrophy”). This search yielded six results (see Appendix A, S1 for list of search terms and also Fig. 1 for the process utilized for our searches).

**Fig 1.**
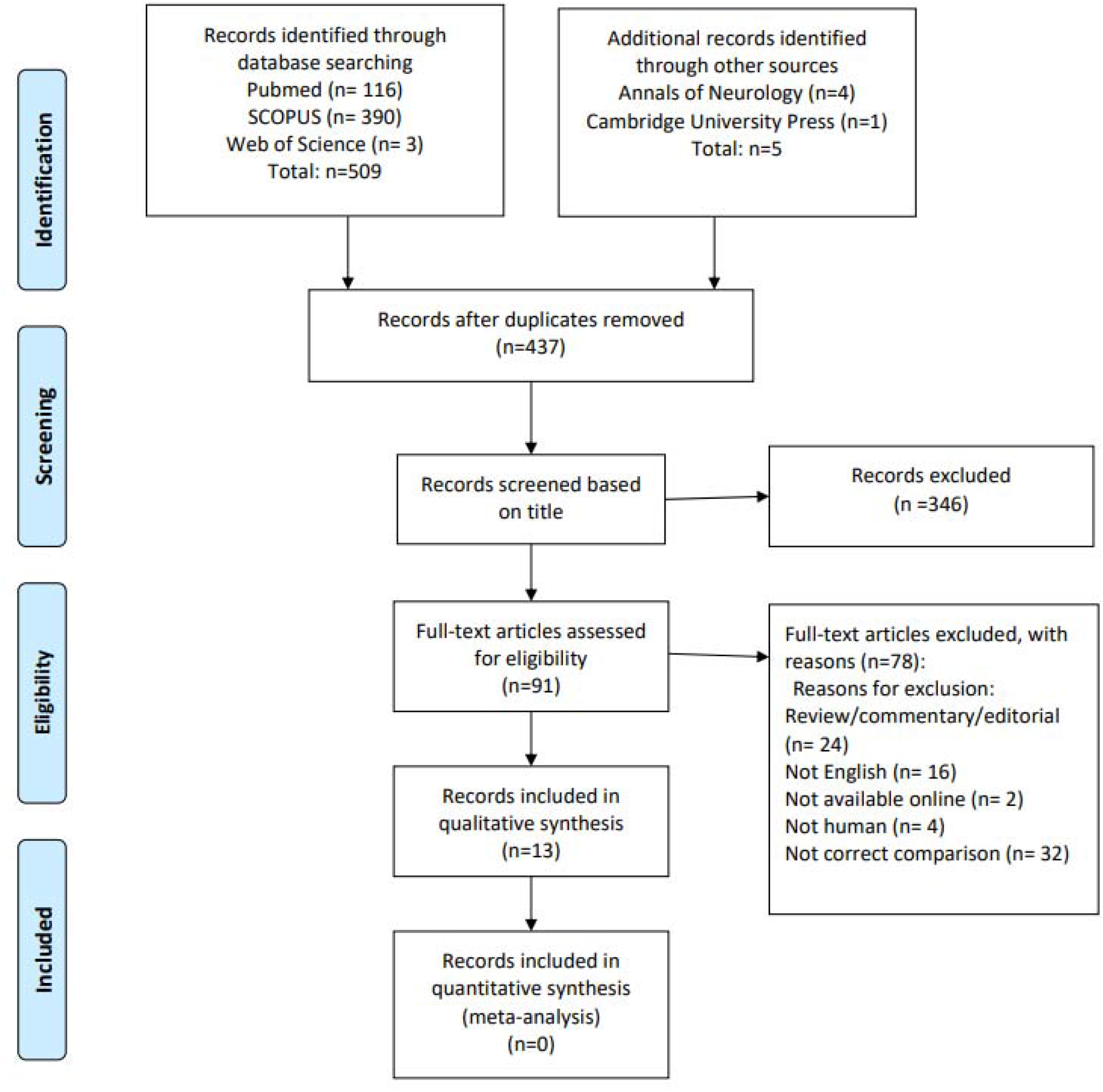
Flow chart of screening, and criteria for inclusion and exclusion of articles (8)

### Study Selection

As SBMA is a relatively rare disease that has not been extensively researched to date, we wished to find as many eligible articles as possible. As such, search filters were not applied during the initial website searches. Results from the PubMed, SCOPUS, and Web of Science database searches, in addition to the Annals of Neurology and Cambridge University Press searches, yielded a total of 515 results (Fig 1, see fig1.docx) that were uploaded to EndNote.

Duplicates were identified and removed using both the duplicate remover function on EndNote, and manually as titles were sorted. This resulted in 346 titles from databases being excluded, with 5 articles identified from individual journal searches also being excluded (Fig. 1). Titles that mentioned SBMA and clinical signs or symptoms and / or pathophysiology, as well as those that mentioned a relationship between CAG repeats and the onset of muscle weakness in male SBMA patients (with or without a female carrier population being part of the study) were included. Ultimately, 91 articles passed the initial title screening process (Fig. 1 and Table 2). Articles were then further screened based on their abstract and full text, which resulted in the exclusion of a further 78 articles. This left a final tally of 13 papers that addressed the aims of this study and which met all of our aforementioned inclusion criteria (Fig. 1).

**Table 2.** Excluded studies from databases based on abstract/full text.

|  |  |
| --- | --- |
| Full text not in English | 16 |
| Did not explicitly compare age of onset of weakness with CAG repeat length in SBMA patients | 32 |
| Review/commentary/editorial paper | 24 |
| Not available online | 2 |
| Not a human study | 4 |
| Total excluded | 78 |

## Results

Our literature search yielded 13 papers eligible for inclusion in this review. The study design of these 13 articles was as follows: ten quantitative observational (7,9–17), one quantitative longitudinal prospective (18), and two quantitative longitudinal retrospective studies (5,19). There were four Japanese studies (5,7,9,12), one German (11), one Canadian (18), one Polish (13), one Taiwanese (19), one Italian (10), two Korean (14,17), one Chinese (16), and one American study (15). Table 4 summarizes the main results of each study. Populations varied depending on the country in which each study took place. Sample sizes ranged between 14 to 223 patients.

**Table 3.**
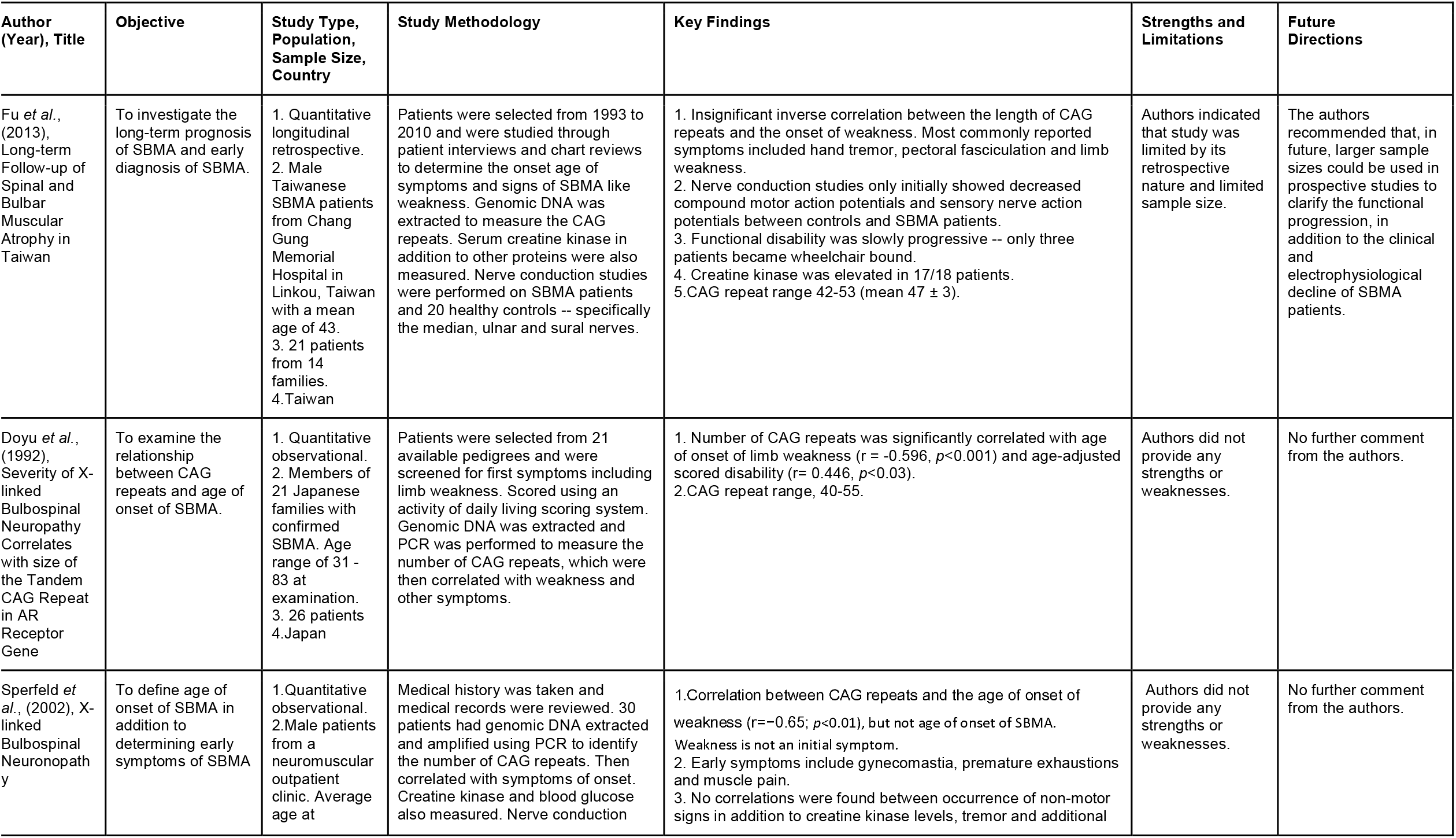
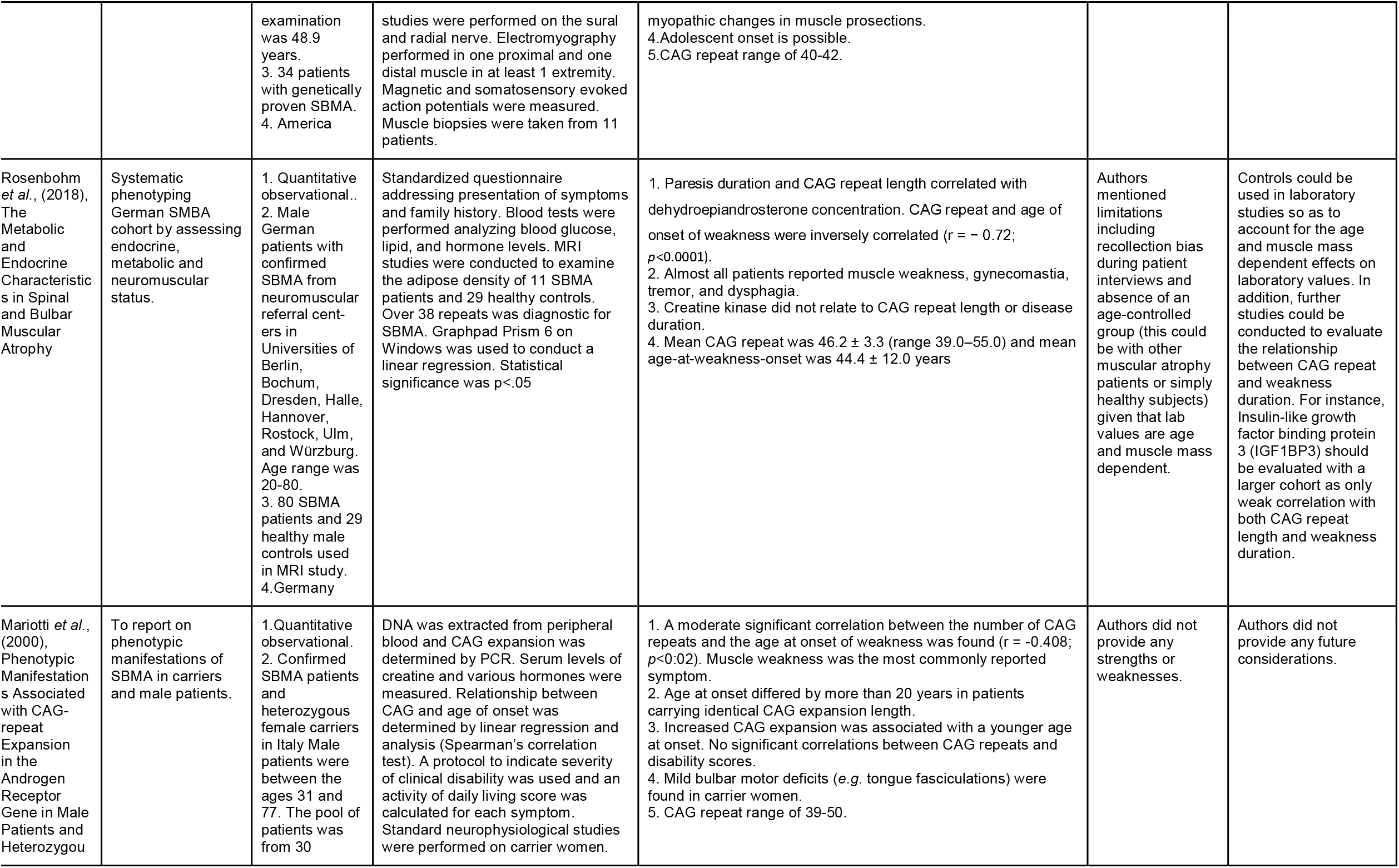
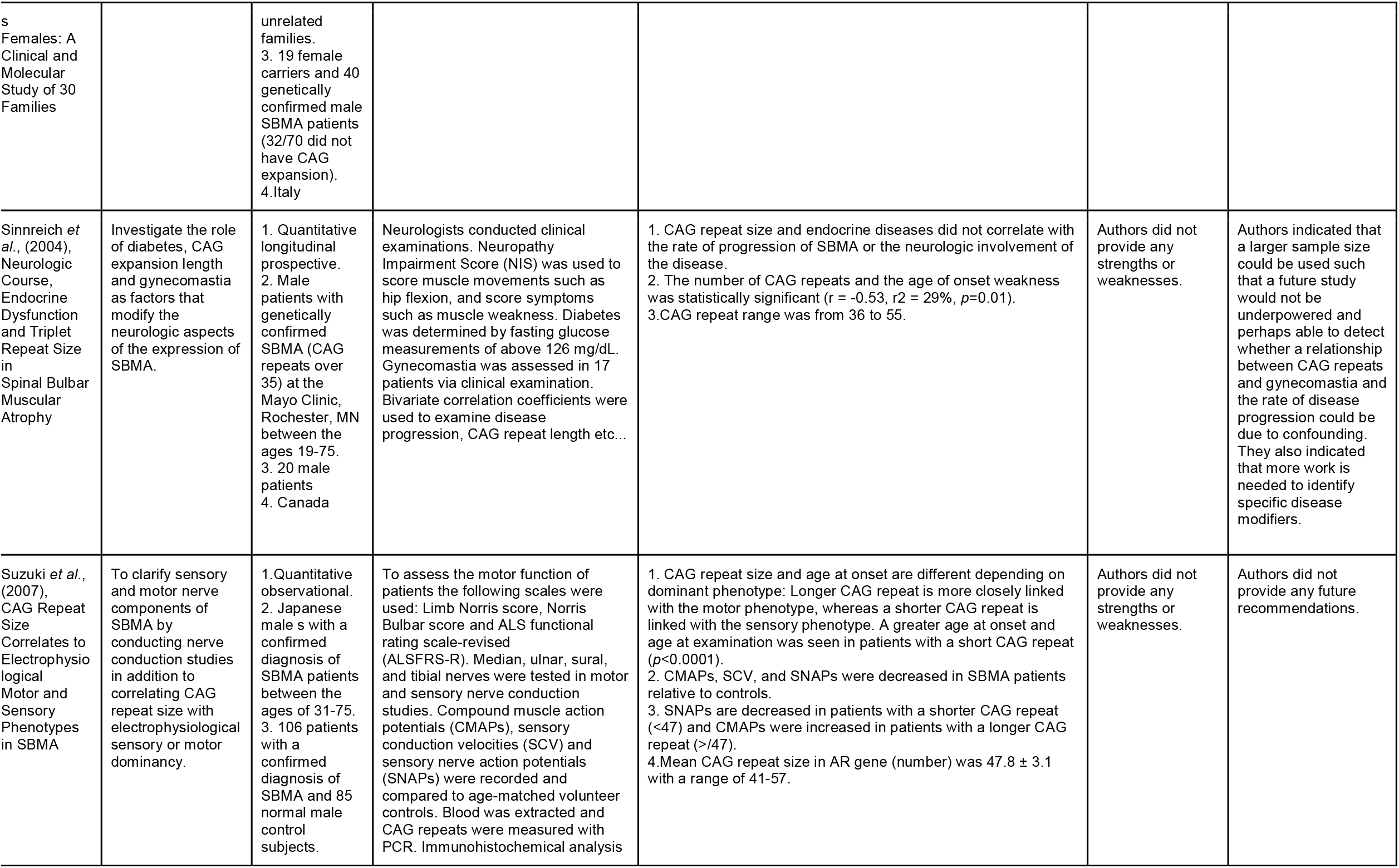
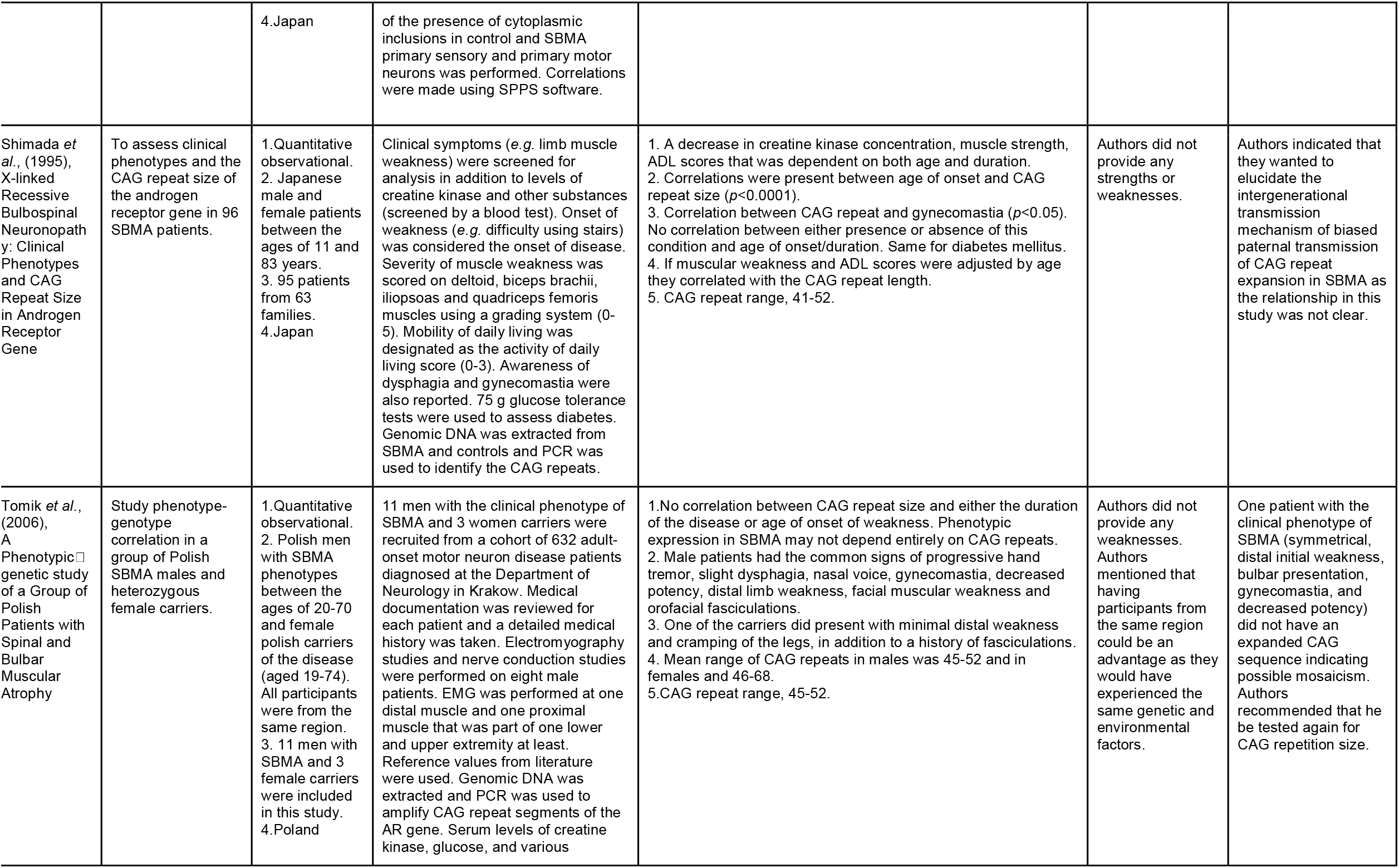
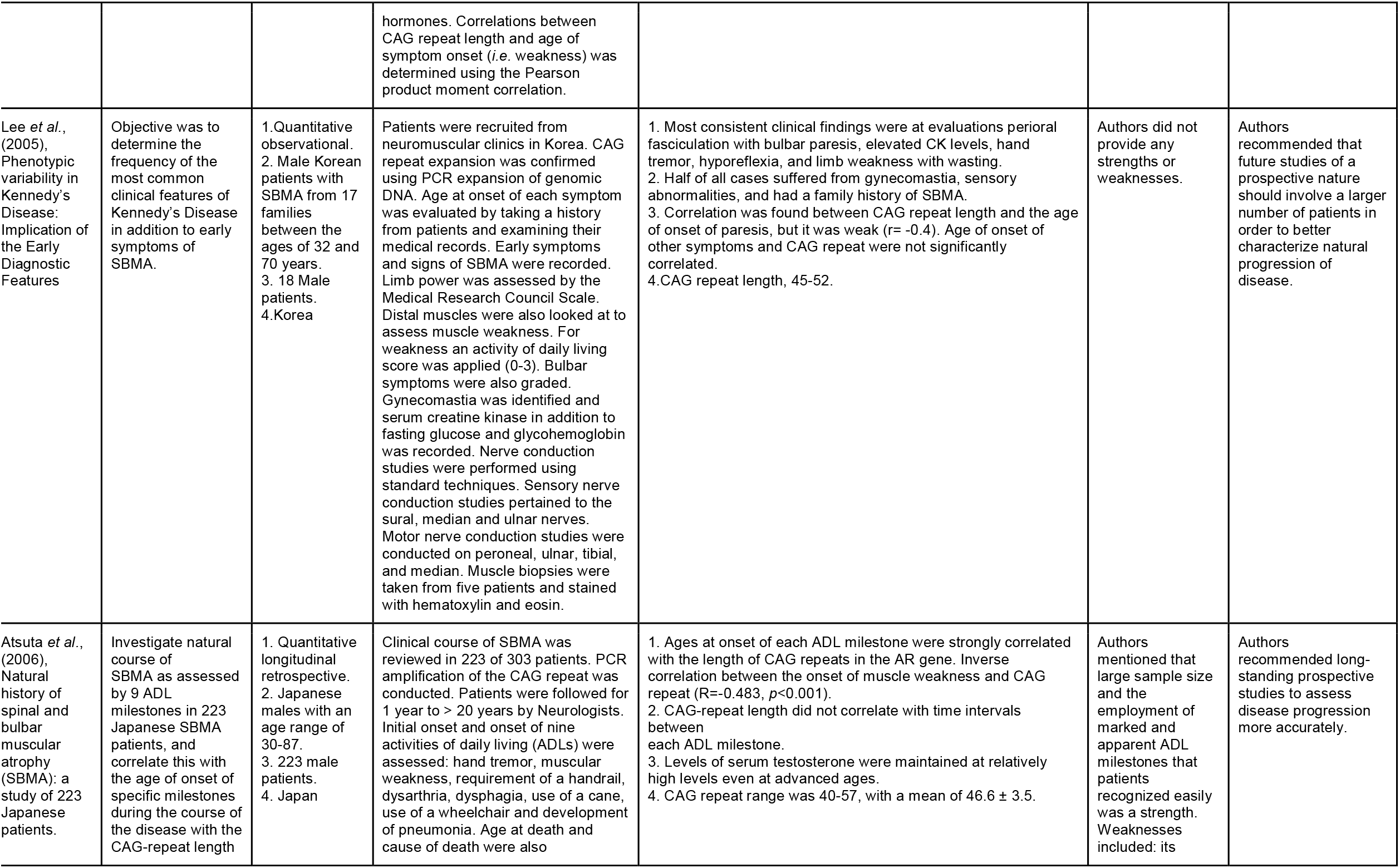
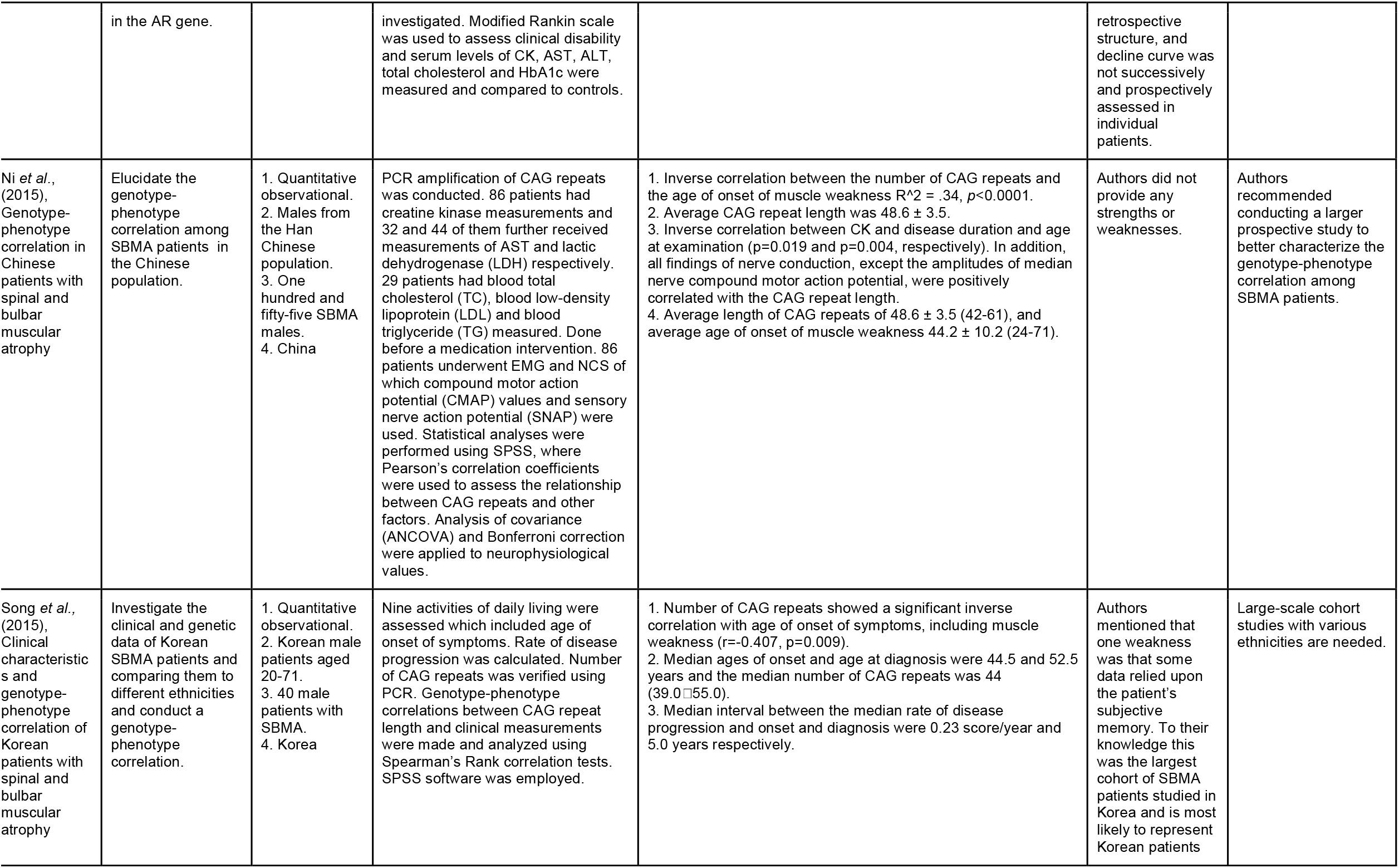
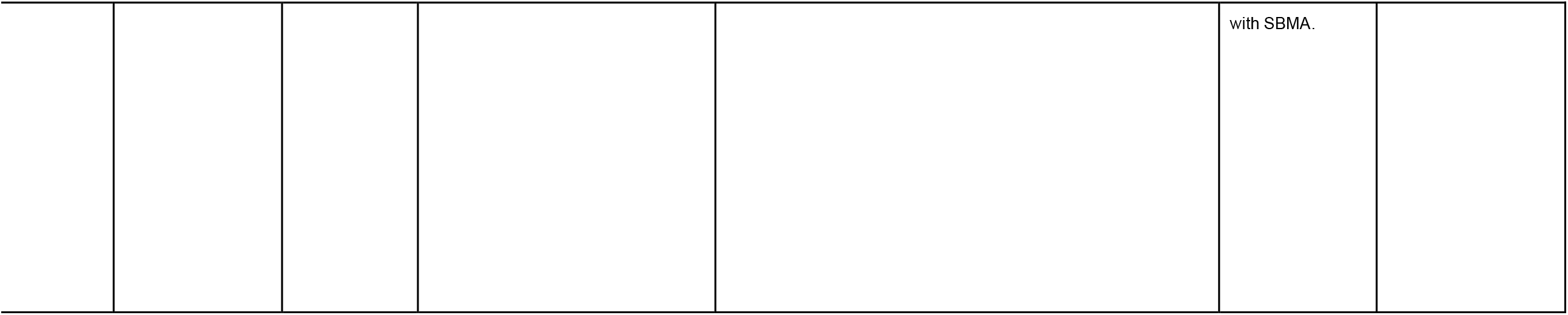
Summary of studies that are included in this review article.

**Table 4.**
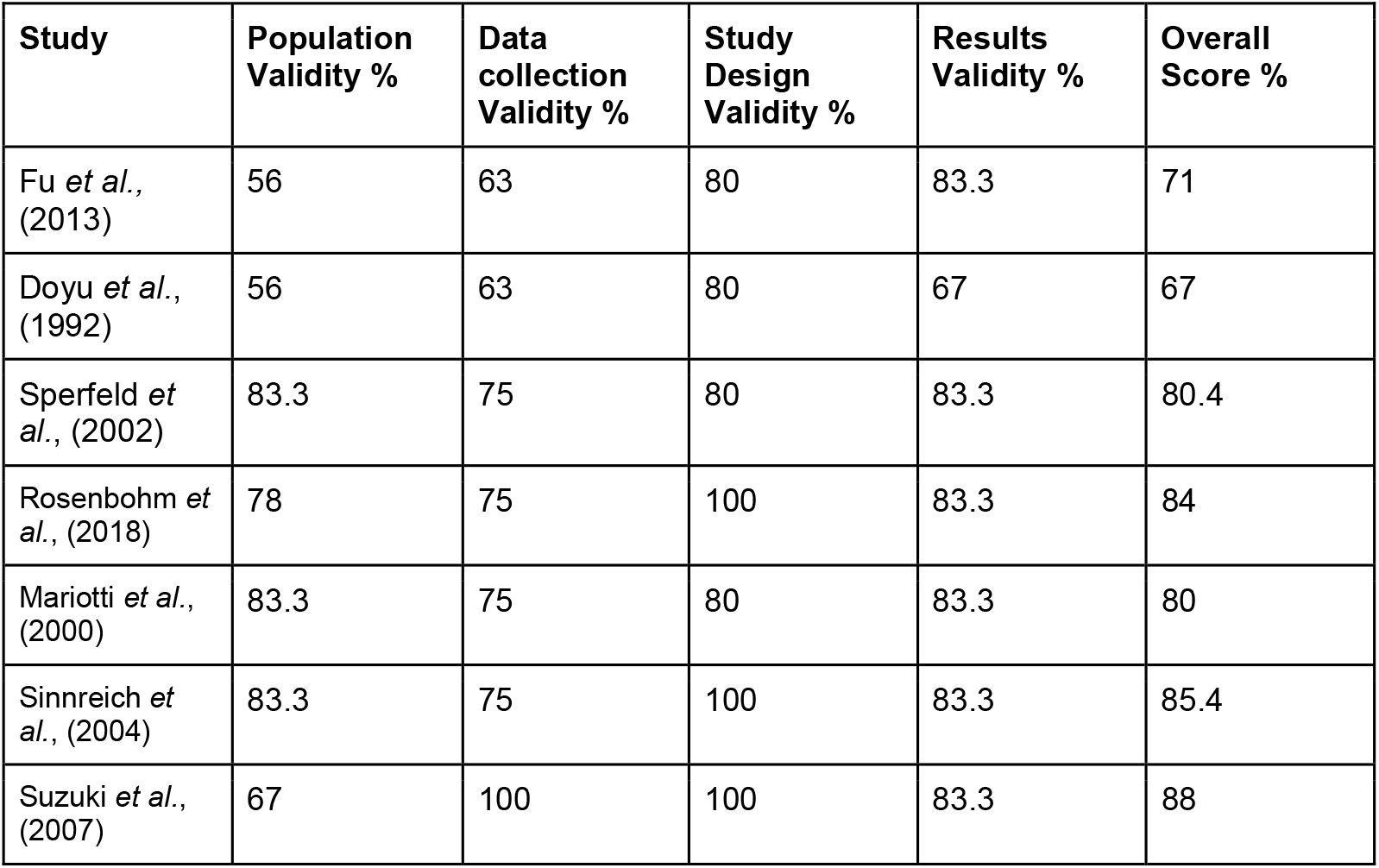
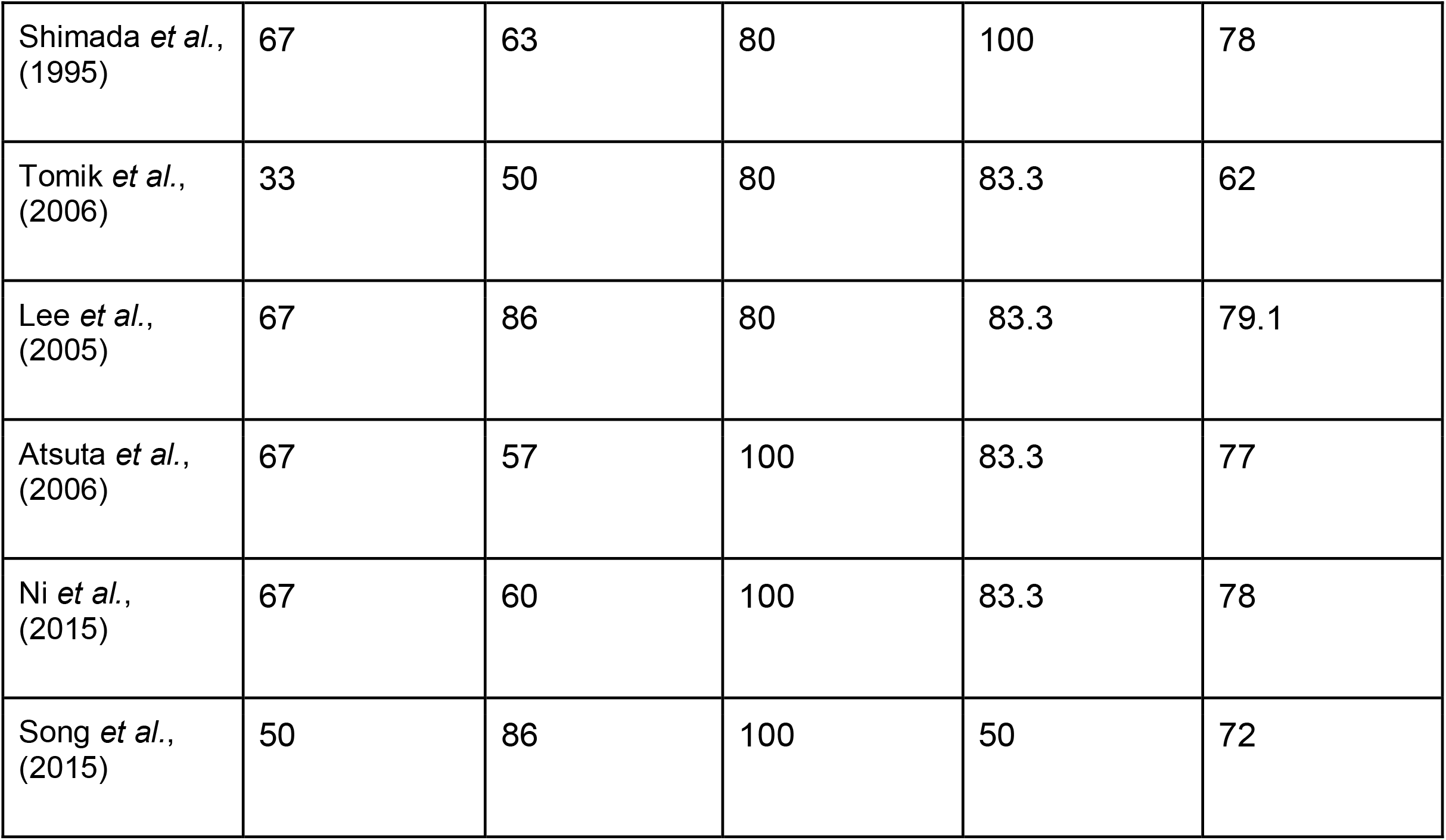
Validity score for quantitative studies using the EBL Critical Appraisal Checklist.

### Critical Appraisal

As all studies were quantitative in nature they were graded using the EBL Critical Appraisal Checklist (20) (see Appendix B, S1 Fig), which provides an overall score for a study based upon the validity of its population, data collection, study design, and results (see Table 4). For a study to be designated as valid, it had to have an overall validity score of 75%. As shown in Table 3, four papers failed this test (9,13,17,19) with overall scores of 67% (Doyu *et al*., 1992), 71% (Fu *et al*., 2013), 72% (Song *et al*., 2015) and 62% (Tomik *et al*., 2006), primarily due to poor population validity, but also due to poor results validity in the cases of Doyu *et al*. (1992) and Song *et al*. (2015) (9,17).

## Objective 1: To explore whether or not the association between CAG repeat number and age of onset of symptoms of SBMA was, or was not, significant

All thirteen eligible studies did examine the relationship between CAG repeat expansion and the age of onset of weakness in SBMA patients (5,7,9–19). These studies could be broadly divided into two groups based on how the authors assessed weakness: studies that used a scoring system to assess weakness and other signs, and studies that used informal patient interviews and/or chart reviews only. Two studies used a combination of methods from both groups (5,14). In the first group, scoring systems included the activity of daily living scores (to assess the onset and severity of weakness) (9,10,14,17), the Limb Norris score, Norris Bulbar score and ALS functional rating scale-revised to assess symptoms and signs of SBMA (12), the Neuropathy Impairment Score to assess motor function and symptoms such as weakness (18), an unspecified standard scoring system to assess symptomatology in patients (11) and a scale to grade weakness (7). The second group of studies simply used either patient interviews (7) or both chart reviews and interviews (13–15,19) or just chart reviews (16) to screen symptoms and/or signs of SBMA.

Studies can be separated into three groups regarding the relationship between CAG repeat length and the age of onset of weakness in SBMA patients (see Table 3). The first group includes articles that found a statistically insignificant inverse correlation (19) and a significant correlation (no r value was provided) (7), or no correlation (13). The second group of studies includes those that revealed either a significant moderate inverse relationship between CAG repeats and the age of onset of weakness (5,9,10,14,16–18) or a strong inverse correlation (11,15). Note that the moderate group also contains results obtained from Lee *et al*. (2005) as, although they claimed a “weak” correlation (r= -0.4) (14), the correlation they identified is actually classified as moderate based on their own published r value (*e.g*., ± 0.1 to ± 0.3 is weak, ± 0.4 to ± 0.6 is moderate, ± 0.7 to ± 0.9 is strong, and ± 1 is perfect) (21). The strength of each group of correlations is based upon accepted r value classification (21). The third group included a study that compared the number of CAG repeats with weakness onset, and related that to either motor-dominant or sensory-dominant phenotypes of weakness. Fewer CAG repeats were correlated with an older age at onset of weakness in SBMA patients and an older age at examination (*p*<0.0001) (12).

## Objective 2: To determine what minimum and maximum number of CAG repeats are associated with weakness onset in SBMA

As shown in Table 4, although all eligible studies did report the number of CAG repeats, the studies could be subdivided into three groups based on the information provided by each article as follows: 1. A group of five studies that listed the ages of participants alongside the minimum and maximum number of recorded CAG repeat number in a retrievable form (7,9,10,14,18); 2. Seven studies that listed CAG repeat number and age ranges (5,11,13,15–17,19); and, 3. One study that involved a unique experimental design, that did not fit into either of the two previous subgroups (12).

In the first group, the article by Sinnreich *et al*. (2004) reported the shortest number of CAG repeats at 36 (weakness onset at age 46) ranging up to 55 (weakness onset at 38), with the cut-off for consideration being any patient with a CAG repeat number greater than 35 (18). Mariotti *et al*. (2000) also reported a minimum CAG repeat number in the thirties, ranging from 39-50, with the patient with the fewest repeats experiencing weakness at age 50 and the two patients with the most repeats experiencing weakness at ages 35 and 31 respectively (10). This study claimed that a normal CAG repeat range was between 12 and 30, and that in SBMA it could be from 39 to 62 (10). There were three studies that reported a minimum number of CAG repeats ≥ 40. Firstly, Doyu *et al*. (1992) who reported a range of 40-55, noting that the patient with the lowest number of repeats had an age of weakness onset of 70, compared to the patient with the most repeats who experienced weakness onset at age 25 (9). Secondly, Shimada *et al*. (1995) who identified a slightly greater minimum repeat number of 41 in a patient who experienced a later age-at-weakness-onset (age 49) than another patient who had the greatest CAG repeat number (52, with the onset of weakness at age 44) (7). Finally, Lee *et al*. (2005) reported the largest minimum CAG repeat number of 45 (age of weakness onset at 55) and a maximum repeat of 54 (onset at 39) (12).

In the second group of studies, the articles by Song *et al*. (2015) and Rosenbohm *et al*. (2018) both reported the lowest minimum CAG repeat number of the group at 39 repeats (11,17). However, their reported age ranges and average CAG repeat number differed, in that Song *et al*. (2015) reported a mean of 44 CAG repeats (range of 39 - 55) and an average age-at-onset of symptoms of SBMA was 44.5 (20.0□71.0) (17), with Rosenbohm *et al*. (2018) reporting a mean age-at-weakness-onset range of 25–75 years (44.4 ± 12.0) and a mean length of CAG expansion of 46.2 ± 3.3 (range 39.0–55.0) (11).

Three of the seven studies in this group reported a minimum number of CAG repeats of exactly 40. Specifically, Sperfeld *et al*. (2002) proposed that the threshold for the number of CAG repeats to be considered as normal is 38, and reported a range of 40-42 CAG repeats among their study population (15). Tomik *et al*. (2006) identified a CAG repeat number range of 45-52, claiming that the number of CAG repeats they considered to be normal was between 5 and 33, but that for SBMA it was between 40 and 62 CAG repeats (13). Atsuta *et al*. (2006) only reported a CAG repeat range of 40-57 for SBMA, with a mean of 46.6 ± 3.5 (5). Finally for this group, the two studies with the highest recorded minimum repeat number were reported in articles by Ni *et al*. (2015), who reported an average number of CAG repeats of 48.6 ± 3.5 (42-61), and an average age of weakness onset of 44.2 ± 10.2 (24-71) years (16), and Fu *et al*. (2013), who found a CAG repeat number range of between 42 and 53 (mean 47 ± 3). They proposed that a normal CAG repeat range is between 14 and 32, suggesting that anything over 40 repeats be regarded as abnormal (19).

One study, by Suzuki *et al*. (2007), did not fit into either group. This study subdivided patients with a CAG repeat size < 47 (short repeats) from those with ≥ 47 (long repeats) and then conducted electromyographic and nerve conduction studies on both groups separately (12). They reported a mean CAG repeat number of 47.8 ± 3.1 with a range of 41-57 repeats (12).

## Discussion

CAG repeat number may account for approximately 60% of clinical heterogeneity in SBMA patients, with genetic, environmental and epigenetic factors also likely to play some role in influencing disease progression (1). However, the contribution of these relative unknowns aside, determining the correlation between the most commonly-mentioned SBMA symptom, weakness, and the primary clinical marker for the condition, CAG repeat number on the AR gene, might potentially uncover a strong indicator of disease onset that could be used to enable early detection, and therefore management, of SBMA. As such, this review is important and timely given that no other review to date has specifically investigated a possible connection between the age of onset of weakness in SBMA and CAG repeat length.

All of the studies investigated for this review addressed our first objective, with eleven of the thirteen indicating that a significant inverse correlation did exist between the CAG repeat number and onset of SBMA weakness (5,7,9–11,15–18) (Table 3). Although the study by Doyu *et al*. (1992) only just failed our initial screening due to poor population validity (56%), data collection validity (63%) and results validity (67%) (overall score 67%) (Table 4) we ultimately included it in this review despite the within-family comparisons which decreased its external validity (9). We felt that the use of disability scores to assess clinical severity in this paper, combined with the limited number of studies available, mitigated against its exclusion. However, notably, a single study did find that shorter CAG repeat number (<47) was associated with an *older* onset of weakness (12).

Only three out of the 13 studies included in this review failed to find a strong or even a moderate statistically significant correlation between CAG repeat number and age of onset of weakness, and these studies often had poor external validity (13,19). For example, the overall validity for the article by Tomik *et al*. (2006) was only 62%, primarily due to its poor population validity (33%; Table 4) (13). Specifically, the lack of age-matched healthy controls to compare EMG results against and the small sample size (11 SMBA patients and 3 carriers; see Table 3) significantly impacted the external validity of the study (13). However, the paper was included as it met the inclusion criteria for this review. Similarly, the study by Fu *et al*. (2013), also had a relatively low overall validity score of 71%, also mostly due to its poor external validity (50%) (Table 4). In this particular case, it was limited by the small population size and retrospective nature (indeed, the authors themselves apparently recognized this deficiency and suggested that any future characterization of the functional progression, and the clinical and electrophysiological aspects of SBMA, be conducted using larger scale prospective studies) (19). The study by Lee *et al*. (2005), which only found what the authors described as a “weak” association (r=0.4) between CAG repeat number and age of onset of weakness in SBMA (14), did pass screening validity with an overall validity score of 78%. However, it is notable that in this case, it too had low population validity (60%; Table 4) and a small sample size which, arguably, affect the conclusions that can be drawn from it. In this case too, the authors recommended that future prospective studies should involve a larger number of patients to better characterize the natural progression of the disease (14).

One final study, which reported only a moderate correlation between CAG repeat length and SBMA weakness (overall score 72%; Table 4), also failed screening due to its poor population and results validity (both 50%) (17). For example, this study had a poor results validity score due to its small sample size and lack of accountability for confounding factors (17). The authors recommended that a large-scale cohort study be conducted (17).

Specifically with regard to our second objective, the majority of studies examined here presented both CAG repeat number data and ages of onset of weakness such that these data could be compared across studies to determine the minimum and maximum number of repeats at age of weakness onset in SBMA (7,9,10,14,18). Rosenbohm *et al*. (2018) and Song *et al*. (2015) simply stated the CAG repeat ranges and the mean age-at-weakness-onset (11,17). Of the five studies that addressed the objective, CAG repeats ranged from the mid-thirties to the mid-fifties, which roughly coincides with the accepted marker of ≥ 38 repeats used to indicate the presence of SBMA (1), and all five studies also found that patients with larger repeat numbers had an earlier onset of weakness (7,9,10,14,18).

### Future Considerations

We believe that any future SBMA studies investigating a possible connection between CAG repeat number and age of weakness onset should account for the potential effects of mosaicism as this may help to explain atypical presentations of SBMA such as one male with only 26 CAG repeats who presented with symptoms of SBMA (*e.g*. distal muscle weakness) (13). Larger sample sizes would also be desirable as this would likely enable a more accurate measurement of the decline in motor function over time (Table 4). Additionally, the use of age-matched controls or at least controls suffering from other muscular atrophies should also be included in studies when conducting electromyographic and/or nerve conduction studies to provide appropriate baseline comparison data. Finally, recollection bias when reporting symptoms could be limited by combining patient interview data, chart reviews and standardized scoring systems in studies so as to minimize the likelihood of any inaccurate or misleading documentation of symptom progression.

### Limitations

One limitation of this review was that it was restricted to papers written in English. This restriction meant that sixteen non-English SBMA articles did not pass the full-text screening process (Fig. 1). Another limitation was the restriction on only including articles that were available online, which meant that two potentially useful studies that could not be accessed online were excluded from our analysis (Fig. 1). Given that very little literature exists on the present topic, four articles that failed screening (9,13,17,19) were included in this review (Table 4). Studies that used only chart reviews and/or patient interviews, but not an objective scoring system to assess symptoms (7,13–16,19) are subject to recollection bias, which may have impacted the recorded age at weakness onset and the correlation of interest.

## Conclusion

Weakness is the most commonly complained of symptom among SBMA patients (1). This review of the relevant SBMA literature has shown that the majority of studies investigating this facet of SBMA pathophysiology have demonstrated an inverse correlation between the number of CAG repeats and the age of weakness onset in SBMA patients. Furthermore, most of the aforementioned studies have proposed that the minimum number of CAG repeats associated with SBMA-linked weakness falls within at least the mid-to-late thirties range or greater. By exploring and revealing the relationship between this reliable marker of SBMA and a common clinical sign of the condition as we have done here, this review not only adds to the relatively scant literature on SBMA, but also, significantly, may facilitate earlier detection of the condition and its subsequent prompt treatment.

## Data Availability

All data produced in the present study are available upon reasonable request to the authors

## Declarations

### Ethics approval

not applicable Consent for publication: not applicable

### Availability of data and materials

The search data analysed during the current study are available from the corresponding author (M.R.) upon reasonable request.

## Author’s Contributions

D.B. – Search, sorting and analysis of data. Author and editor of article. M.R. – Corresponding author, check of search and data analysis, and editor, of article.

## Acknowledgements

none

## Supporting Information

S1 File. Appendix A search terms.

S1 Fig. Appendix B EBL data.

