## Supplementary figures and images for "A Systematic Review of the Association between the Age of Onset of Spinal-Bulbar Muscular Atrophy (Kennedy’s Disease) and the Length of CAG Repeats in the Androgen Receptor Gene"

### suppl fig 1

**Appendix B**

EBL checklists

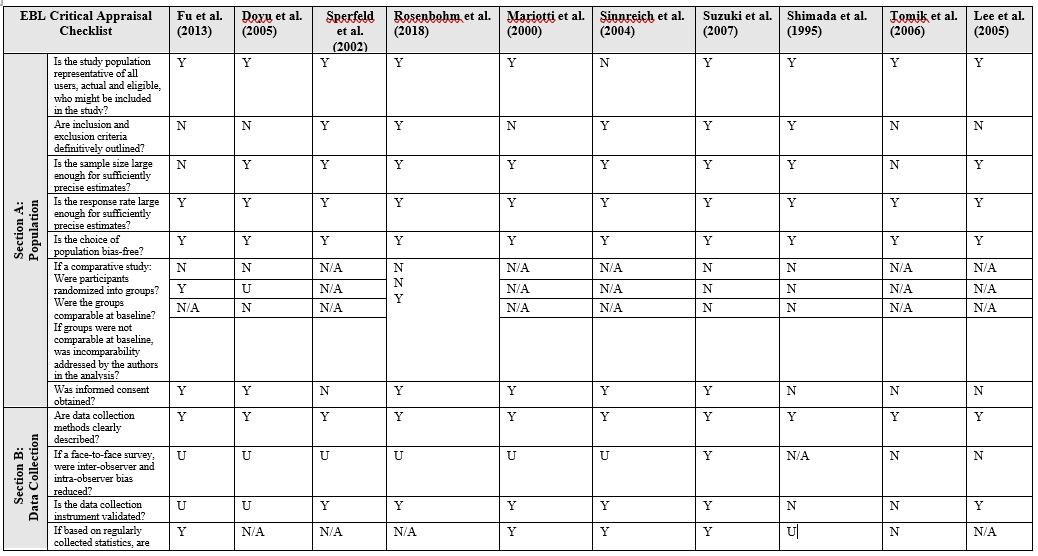

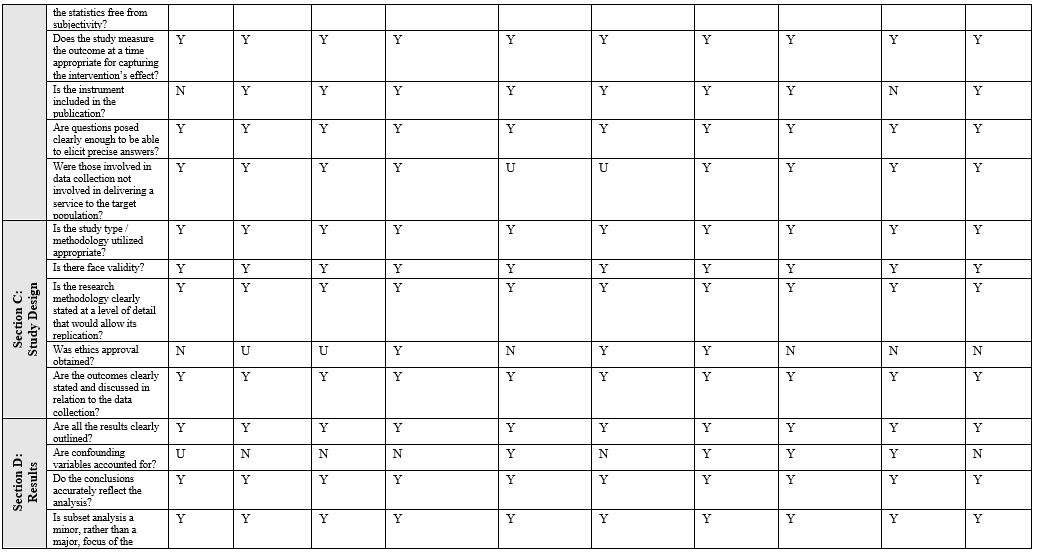

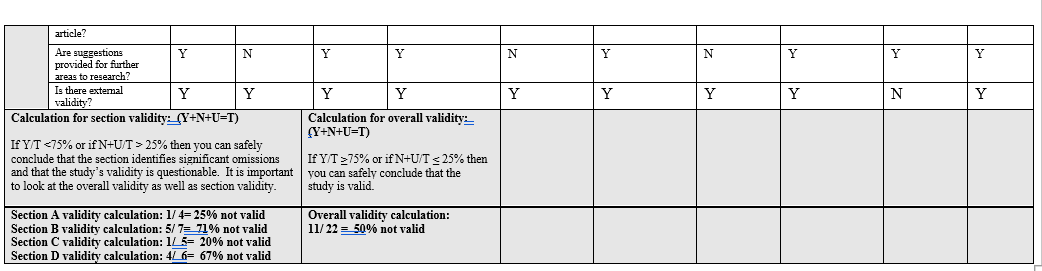

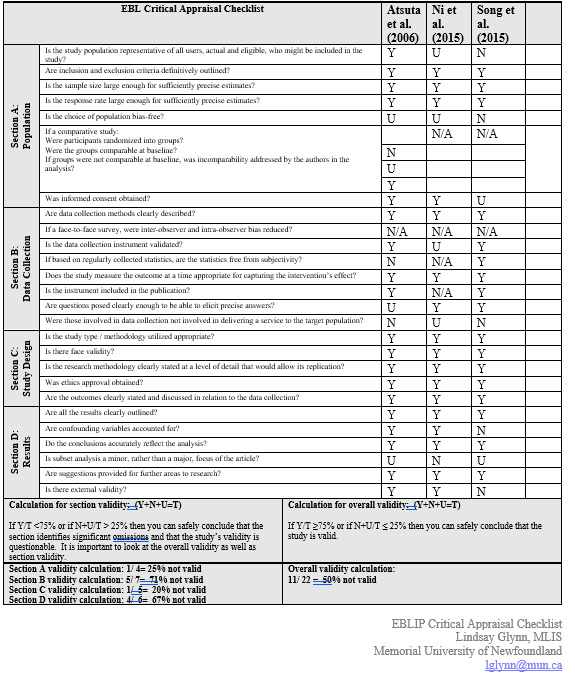
