## Supplementary material for "A Systematic Review of the Association between the Age of Onset of Spinal-Bulbar Muscular Atrophy (Kennedy’s Disease) and the Length of CAG Repeats in the Androgen Receptor Gene": suppl file 1

**Appendix A**

**Pubmed (MEDLINE) search terms:**

("Bulbo-Spinal Atrophy, X-Linked" [Mesh] OR “spinal bulbar muscular atrophy”[tw] OR “kennedy’s disease”[tw] OR “bulbospinal muscular atrophy”[tw]) AND (( "Muscle Weakness/diagnosis"[Mesh] OR "Muscle Weakness/etiology"[Mesh] OR "Muscle Weakness/genetics"[Mesh] OR "Muscle Weakness/pathology"[Mesh] OR "Muscle Weakness/physiology"[Mesh] OR "Muscle Weakness/physiopathology"[Mesh] OR "Muscle Weakness/prevention and control"[Mesh] ) OR "muscle weakness"[tw] OR weak*[tw] OR paresis[tw] OR symptom*[tw]) AND ("age of onset"[tw] OR onset[tw] OR "age at onset"[tw]) AND ("Trinucleotide Repeat Expansion"[Mesh] OR "CAG repeat"[tw]) NOT (Hunting*[tw] OR "Huntington's disease"[tw] OR "Fragile X"[tw] OR "myotonic dystrophy"[tw] OR "amyotrophic lateral sclerosis"[tw] OR "ALS"[tw] OR "spinocerebellar ataxia"[tw] OR "oculopharyngeal muscular dystrophy"[tw]) .

**Web of Science search terms**

("Bulbo-Spinal Atrophy, X-Linked" OR "spinal bulbar muscular atrophy" OR "kennedy’s disease" OR "bulbospinal muscular atrophy") AND (“Muscle weakness” OR weak* OR paresis OR symptom*) AND ("age of onset" OR onset) AND ("Trinucleotide Repeat Expansion" OR "CAG repeat")

**SCOPUS Search terms**

( ALL ( ( "Bulbo-Spinal Atrophy, X-Linked"  OR  "spinal bulbar muscular atrophy"  OR  "kennedy's disease"  OR  "bulbospinal muscular atrophy" )  AND  ( "Muscle weakness"  OR  weak*  OR  paresis  OR  symptom* )  AND  ( "age of onset"  OR  onset )  AND  ( "Trinucleotide Repeat Expansion"  OR  "CAG repeat" ) ) )

**Cambridge University Press search terms**

“SBMA”

**Annals of Neurology - Wiley Online Library search terms**

"Manabu Doyu" AND "spinal bulbar muscular atrophy"
